# Pentavalent Vaccine: How Safe Is It Among Infants Accessing Immunization In Nigerian Health Facilities

**DOI:** 10.1101/2024.05.28.24307998

**Authors:** Elemuwa Uchenna Geraldine, Enato Ehijie, Elemuwa Christopher Ononiwu, Elemuwa Tochukwu Daniel, Morufu Olalekan Raimi

## Abstract

**Rationale:** Pentavalent vaccines offer significant public health benefits by protecting against five major diseases with a single injection. However, concerns have been raised in various studies regarding potential associations between combined vaccines and conditions such as autism, febrile seizures, sudden unexpected death in infancy, demyelinating disorders, and neurodevelopmental disorders.

**Objective:** This study aimed to evaluate the safety of pentavalent vaccines administered to infants aged between 6 and 14 weeks.

**Methods:** A total of 423 infants, all aged 6 weeks and receiving their first pentavalent vaccine at selected healthcare facilities, were recruited for the study after obtaining informed consent from their mothers or caregivers. The infants were administered three doses of the vaccine at 6, 10, and 14 weeks. Mothers and caregivers were provided with diaries and thermometers to monitor and record any Adverse Events Following Immunization (AEFI) observed in their babies after each vaccine dose. Follow-up was conducted through telephone calls to ensure accurate monitoring and recording of any identified events.

**Results:** The study identified various AEFIs in the infants following their routine immunizations. These included pain at the injection site, fever, swelling at the injection site, vomiting, refusal to feed, excessive crying, coughing, rash, stooling, restlessness, and severe local reactions. Fever was the most commonly reported systemic AEFI, with incidence rates decreasing from 66.98% after the first dose to 55.37% after the third dose. Pain and swelling at the injection site were the most frequently reported local AEFIs, with their incidence also decreasing from the first to the third doses. No statistically significant differences were observed in the occurrence of AEFIs across the three doses.

**Conclusions:** The pentavalent vaccine was found to be safe for infants in the Federal Capital Territory (FCT), Nigeria, with the observed AEFIs being generally mild and decreasing in frequency with subsequent doses.

**Recommendations:** Further studies should be conducted to monitor long-term safety and potential rare adverse effects of pentavalent vaccines. Additionally, public health education should emphasize the safety and benefits of pentavalent vaccines to increase vaccination rates and reduce vaccine hesitancy.

**Significance Statement:** This study underscores the safety of pentavalent vaccines in infants, reinforcing their role in preventing multiple serious diseases through a single immunization schedule. The findings support the continued use and promotion of pentavalent vaccines in public health programs, particularly in regions with high infant mortality rates and limited healthcare resources.

## 1. Introduction

A combination vaccine, as used in this context, refers to a single vaccine formulation that contains multiple antigens, each of which confers immunity against different vaccine-preventable diseases [1, 2]. Combination vaccines are particularly advantageous in addressing the complexities of routine childhood immunization programs. They streamline the vaccination process by reducing the number of injections required, which improves compliance and increases immunization uptake among infants and their parents. This efficiency also allows for the inclusion of additional vaccines in the already crowded immunization schedules, enhancing overall public health outcomes [3–5]. Key examples of combination vaccines include the measles/mumps/rubella (MMR) vaccine, which protects against three diseases, and tetravalent vaccines such as DTP-Hib (diphtheria, tetanus, pertussis, and Haemophilus influenzae type b) or DTP-HepB (diphtheria, tetanus, pertussis, and hepatitis B) [6–16]. The pentavalent vaccine (DTP-HepB-Hib) further expands this protection by covering five diseases in a single injection. Despite extensive safety data collected for the individual antigens used in these combination vaccines during both pre-licensure and post-licensure clinical assessments, the potential for new safety issues emerging from their combined use cannot be completely ruled out. Continuous and rigorous safety monitoring is essential to ensure that the benefits of combination vaccines continue to outweigh any potential risks [17–19]. The necessity for comprehensive vaccine safety monitoring cannot be overstated. This monitoring should encompass a holistic approach, considering human factors, technical capabilities, as well as planning and management aspects within our society [20–25]. Such an inclusive perspective ensures that vaccine safety assessments are thorough and that any emerging safety concerns are promptly identified and addressed, maintaining public trust in immunization programs [8, 11, 12, 14, 21–25].

Hence, the pentavalent vaccine, a combination vaccine, was introduced into Nigeria’s routine immunization schedule in 2012, replacing the previously used DPT vaccine. This vaccine has since become a cornerstone of routine immunization efforts, providing protection against five critical childhood diseases: *Haemophilus influenzae type B* (which can cause meningitis, pneumonia, and otitis), whooping cough (pertussis, caused by Bordetella pertussis), tetanus, hepatitis B, and diphtheria [1–3]. During the initial rollout of the pentavalent vaccine in various countries, several safety concerns were identified. While extensive studies have thoroughly assessed the safety of the individual antigens included in the pentavalent vaccine, the potential for synergistic effects when these antigens are combined cannot be completely dismissed. Continuous monitoring and evaluation are necessary to ensure the ongoing safety and efficacy of the vaccine [19–25]. According to the World Health Organization (WHO), Adverse Events Following Immunization (AEFIs) are defined as any untoward medical occurrences that follow immunization, which may or may not be causally related to the vaccine. AEFIs can range from serious to non-serious and may result from various factors, including vaccine-related effects, defects in vaccine quality, errors in the immunization process, anxiety-related reactions, or coincidental events that are not related to the vaccine itself [26]. Understanding and addressing these AEFIs is crucial for maintaining public confidence in vaccination programs. Comprehensive safety monitoring helps to identify and manage any adverse events, ensuring that the benefits of immunization continue to outweigh the risks, and contributing to the overall success of public health initiatives [4–9].

During the initial introduction of the pentavalent vaccine, significant safety concerns emerged in four Asian countries: Sri Lanka, Bhutan, India, and Vietnam. Reports surfaced of suspected hypotonic-hyporesponsive episodes (HHE), encephalopathy, and/or meningoencephalitis, and there were some recorded deaths among vaccinated infants. These incidents prompted immediate attention due to the robust safety monitoring systems in place in these countries, which facilitated thorough causality assessments of the suspected vaccine-event combinations [17–23]. The comprehensive investigations conducted in these countries concluded that the pentavalent vaccine was not responsible for the identified adverse events. This finding was crucial as it allowed for the reintroduction of the previously suspended pentavalent vaccine into their routine immunization schedules, reaffirming the vaccine’s safety [20–25].

Furthermore, as the pentavalent vaccine was still relatively new during its early introduction phase, ongoing research was essential to continually evaluate its safety profile [20, 21, 22, 27]. For instance, Sreelakshmi *et al.* [28] conducted a study on the effectiveness and impact of the pentavalent vaccination program in India and other South Asian countries. Their study reported the deaths of approximately 100 otherwise healthy infants. Although the researchers noted that these deaths were not necessarily causally related to the vaccine, they emphasized that the potential for unknown attributes of the vaccine could not be completely dismissed [22–26]. In light of these findings, continuous and rigorous monitoring of the pentavalent vaccine’s safety in various settings and environments was recommended. This ongoing surveillance is crucial to adequately characterize the vaccine’s safety profile, ensuring that any potential adverse effects are promptly identified and managed, thereby maintaining public trust in immunization programs and ensuring the sustained success of these critical public health initiatives [6, 8, 9, 13, 14, 28]. Eregowda *et al.* [29] conducted a study to evaluate the immunogenicity and tolerability of the pentavalent vaccine, Quinvaxem®. The safety aspect of this study involved monitoring solicited local and systemic reactions, with mothers of the participating infants provided with diaries to record any Adverse Events Following Immunization (AEFIs) observed in their children. The findings indicated that the vaccine was well-tolerated, with no serious adverse events (AEs) attributable to the vaccine [29].

Several other researchers have also investigated the safety of pentavalent vaccines in diverse settings. Bavdekar *et al.* [30], Angela *et al.* [31], Dodoo *et al.* [32], Sharma *et al.* [33], Asturias *et al.* [34], Maria *et al.* [35], and Manoochehr *et al.* [36] conducted studies across various populations. These studies predominantly identified non-serious AEFIs, which typically resolved with or without medication. Furthermore, the frequency of these AEFIs generally decreased across the three doses of the pentavalent vaccine [30–36]. To specifically characterize the safety profile of the pentavalent vaccine within the Nigerian population, an intensive prospective study was conducted. This study aimed to assess the safety and tolerability of the pentavalent vaccines introduced into Nigeria’s routine immunization program [1, 2]. Through detailed monitoring and analysis, this study provided crucial data to support the continued use of the pentavalent vaccine, ensuring its benefits far outweigh any potential risks and thereby reinforcing public health initiatives.

## 2. Materials and Methods

### 2.1 Study Design

A prospective and observational active monitoring method was implemented to evaluate and document Adverse Events Following Immunization (AEFIs) associated with pentavalent vaccines. This study focused on infants aged between 6 and 14 weeks who were immunized according to the approved routine immunization schedule in Nigeria. The infants were monitored in real-life conditions to ensure that the findings accurately reflected the vaccine’s performance and safety in everyday clinical settings. This approach allowed for a comprehensive assessment of the safety profile of the pentavalent vaccine under typical use, providing valuable data to inform public health strategies and vaccination policies.

### 2.2 Study Location

A stratified random sampling method was employed to select study sites across the six Area Councils of Abuja, the federal capital city of Nigeria. Health facilities within these councils were categorized into three levels: primary, secondary, and tertiary. From each category, a random sampling of health facilities was performed to ensure a representative selection. The study was conducted in a total of fourteen health facilities, encompassing a diverse range of healthcare settings. These facilities included: University Teaching Hospital, Gwagwalada, National Hospital Abuja, Asokoro General Hospital, Abaji General Hospital, Kwali General Hospital, Kubwa General Hospital, Nyanya General Hospital, Kuje General Hospital, PHC Clinic Dutse Makarantha, PHC Dabi Bako Kwali, PHC Kuje, PHC Nuku Abaji, PHC T/Maje Gwagwalada and PHC Idu. This comprehensive selection ensured that the study covered a broad spectrum of healthcare environments, from highly specialized tertiary hospitals to primary health care centers, thereby providing a holistic view of the pentavalent vaccine’s safety and tolerability across different levels of the healthcare system in Abuja.

### 2.3 Study Population

The study population for evaluating the safety of pentavalent vaccines consisted of 6-week-old infants receiving their first dose of the pentavalent vaccine.

### Inclusion Criteria

i. Infants receiving their first immunization with the pentavalent vaccine.
ii. Infants aged exactly 6 weeks at the time of the first dose.
iii. Infants receiving their immunization exclusively at the designated health facilities where the study was conducted.
iv. Infants whose parents or guardians provided informed consent for participation in the study.

### Exclusion Criteria

i. Infants experiencing acute illness at the time of enrollment, as this could confound the identification of vaccine-related adverse events due to overlapping symptoms of the illness or other underlying conditions.
ii. Infants who had either completed their immunization schedule or initiated their first dose of the pentavalent vaccine at a different health facility outside the study sites.

By adhering to these criteria, the study ensured a consistent and specific population for accurately assessing the safety and tolerability of the pentavalent vaccine within the selected health facilities.

### 2.4 Sample Size

The sample size for monitoring children receiving the pentavalent vaccine was determined using a prevalence rate of 50% for Adverse Events Following Immunization (AEFIs). The calculation employed the method for determining sample size for a prevalence survey with finite population correction, as outlined by Philippe Glaziou [37]. *Assumptions and Parameters:*

`o` Precision: 5%
`o` Prevalence: 50%
`o` Population of children under 1 year in the Federal Capital Territory (FCT): 125,135.3
`o` Confidence Interval: 95%, with specified limits of 45% to 55% (equal to prevalence plus or minus precision)

Using these parameters, the estimated sample size was calculated to be 385 infants. To account for potential non-response or drop-out, a 10% non-response rate was added to the initial estimate, resulting in an adjusted sample size of 423 infants. This approach ensured a robust and reliable sample for evaluating the safety of the pentavalent vaccine in the target population.

### 2.5 Data Collection Tool

All 6-week-old infants who visited the selected health facilities during the study period were evaluated based on the inclusion criteria. Recruitment occurred after mothers provided informed consent to participate. Mothers were given the opportunity to ask questions, and for those who could not read, the study procedures were explained in a language they understood. Once a mother comprehended the study requirements and agreed to participate by signing the informed consent form, her child was enrolled in the study. For illiterate mothers who could not read or write but expressed a willingness to participate, a literate person living with them was invited to receive training on the study requirements. The infants were monitored over a three-month period, coinciding with the administration of the three doses of the pentavalent vaccine at 6 weeks, 10 weeks, and 14 weeks. Additionally, the participants received the oral poliovirus vaccine as per the routine immunization schedule.

Each mother received a diary, formatted as an exercise book, which included the child’s demographic information (name, date of birth, state of origin, and date of immunization) and instructions for monitoring temperature at specified intervals. Mothers were trained to identify and record Adverse Events Following Immunization (AEFIs) in their infants and were provided with digital thermometers for this purpose. Mothers were instructed to monitor their babies closely for the first three days after each immunization and to document any adverse events observed. They were also advised to continue recording any new events until the next immunization appointment. Follow-up phone calls by the researchers and their assistants ensured that mothers were consistently monitoring and documenting observations. Mothers were encouraged to bring their infants to the health facilities for reassessment and treatment by a doctor if any serious adverse events occurred.

At each subsequent immunization clinic visit, the diaries were reviewed and returned to the mothers for ongoing monitoring and recording. After the infants received their third pentavalent vaccination and the monitoring period concluded, mothers were required to submit the diaries during their next clinic visit. All adverse events (both serious and non-serious) experienced by the infants during the study period, as recorded by the mothers or caregivers, were collected and analyzed from the diaries. This detailed approach ensured comprehensive and accurate monitoring of the infants, facilitating a thorough assessment of the pentavalent vaccine’s safety profile in the study population.

### 2.6 Study variables

The outcome variables for this study encompassed all reported adverse events following vaccination, categorized into local and systemic reactions.

### Local Reactions

`o` **Pain at the injection site**: Discomfort or pain localized to the area where the vaccine was administered.
`o` **Swelling at the injection site**: Inflammation or swelling in the area of the injection.
`o` **Severe local reactions**: Intense localized responses potentially including significant swelling, redness, or hardening of the injection site.
`o` **Generalized rashes**: Widespread skin rashes appearing post-vaccination.

### Systemic Reactions

`o` **Fever**: Elevated body temperature following vaccination.
`o` **Persistent crying**: Continuous crying that is prolonged and beyond typical crying durations.
`o` **Vomiting**: Episodes of throwing up post-vaccination.
`o` **Stooling**: Changes in bowel movements, potentially including diarrhea.
`o` **Shock**: Severe, life-threatening reaction characterized by rapid onset of symptoms like difficulty breathing, low blood pressure, and loss of consciousness.
`o` **Restlessness**: Increased irritability and inability to stay calm or still.
`o` **Refusal to suck**: Infants not feeding or sucking as usual.
`o` **Coughing**: Episodes of cough observed after immunization.

These variables provided a comprehensive framework for documenting and analyzing the range of adverse events experienced by infants’ post-vaccination, thereby contributing to a thorough assessment of the pentavalent vaccine’s safety profile.

### 2.7 Data Analysis

Descriptive statistics, employing frequencies and percentages, were utilized to characterize the socio-demographic attributes of infants whose mothers submitted their diaries. We further investigated bivariate relationships between the infants’ sex and each adverse reaction using the chi-square test, aiming to discern any potential associations. The reported Adverse Events Following Immunization (AEFIs) were systematically entered and analyzed using the Statistical Package for Social Sciences (IBM® SPSS) Version 23. Statistical significance was assessed at a significance level of P ≤ 0.05, ensuring rigorous evaluation of any observed associations or patterns between adverse reactions and infant characteristics. This analytical approach facilitated a comprehensive examination of the data, enabling us to identify potential correlations between the infants’ sex and adverse reactions, thereby contributing valuable insights into the safety profile of the pentavalent vaccine within the study population.

## 3. Results

A total of 423 infants were initially enrolled in the study, with data received from 235 (55.5%) infants whose mothers submitted their babies’ diaries for final analysis. This sample size was deemed sufficient for drawing conclusions regarding the safety of pentavalent vaccines, allowing for comparisons with findings from other studies. Various reasons were provided for the loss to follow-up among some mothers whose infants were recruited for pentavalent vaccine monitoring. These reasons included:

- Relocation to other states or area councils not covered by the study.
- Completion of the infant’s immunization at health facilities not included in the selected monitoring sites.
- Some mothers perceived no necessity for continued monitoring, while others simply reported that their infants were healthy and exhibited no adverse reactions.

Among the 235 infants who completed pentavalent monitoring, 112 (47.70%) were male, and 123 (52.30%) were female. Data were received from infants recruited from nine out of the fourteen health facilities, representing primary, secondary, and tertiary institutions across the six Area Councils of Abuja FCT, categorized into urban and rural locations (Table 1 and figure 1 below). Analysis revealed no statistically significant differences in adverse event occurrence based on the infants’ sex. This finding underscores the importance of considering various factors in assessing adverse event occurrences and highlights the need for continued monitoring to ensure the safety of pentavalent vaccines across diverse populations and settings (Table 2).

**Fig 1:**
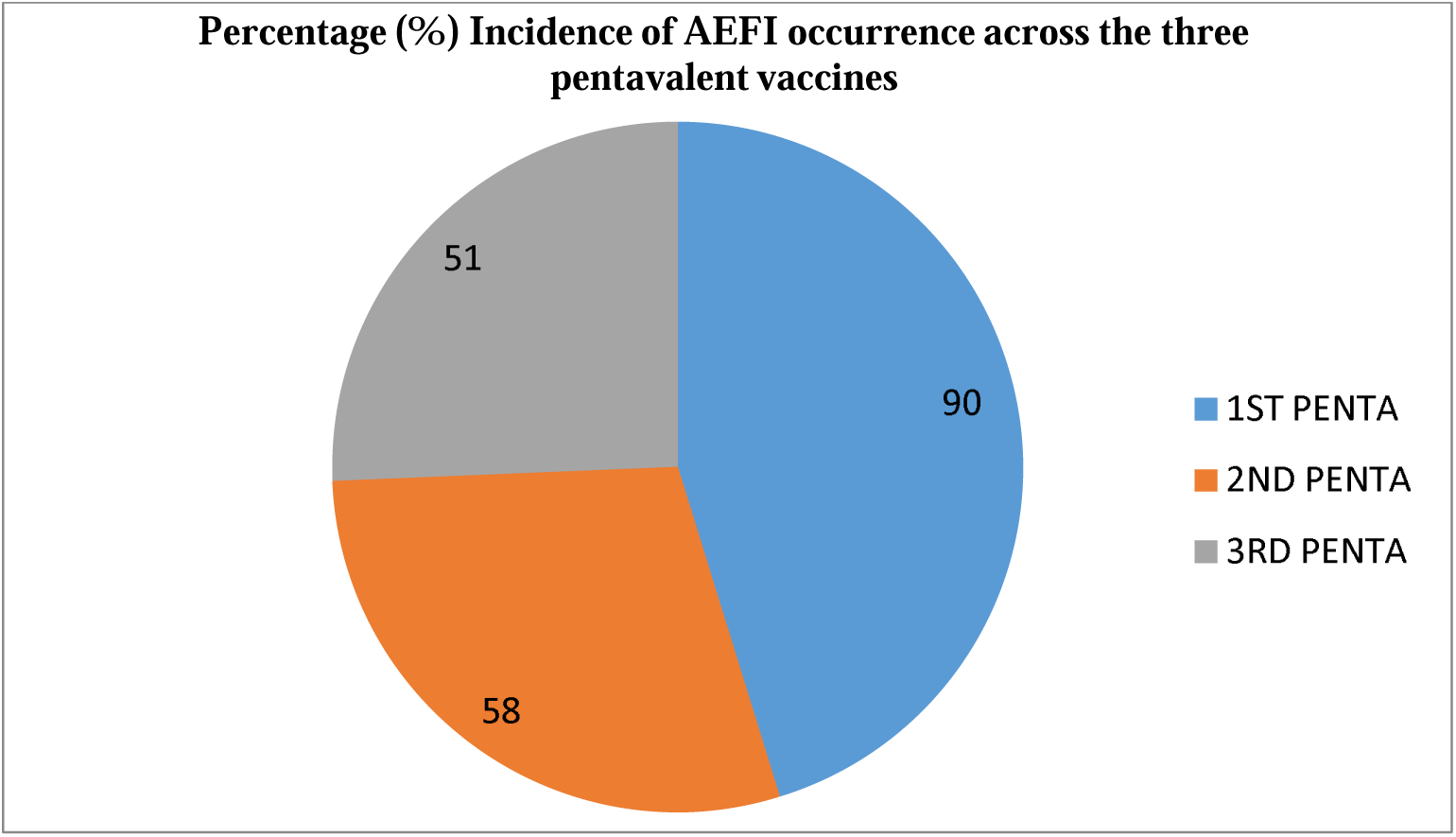
Incidence of AEFI occurrence across the three pentavalent vaccines.

**Table1:**
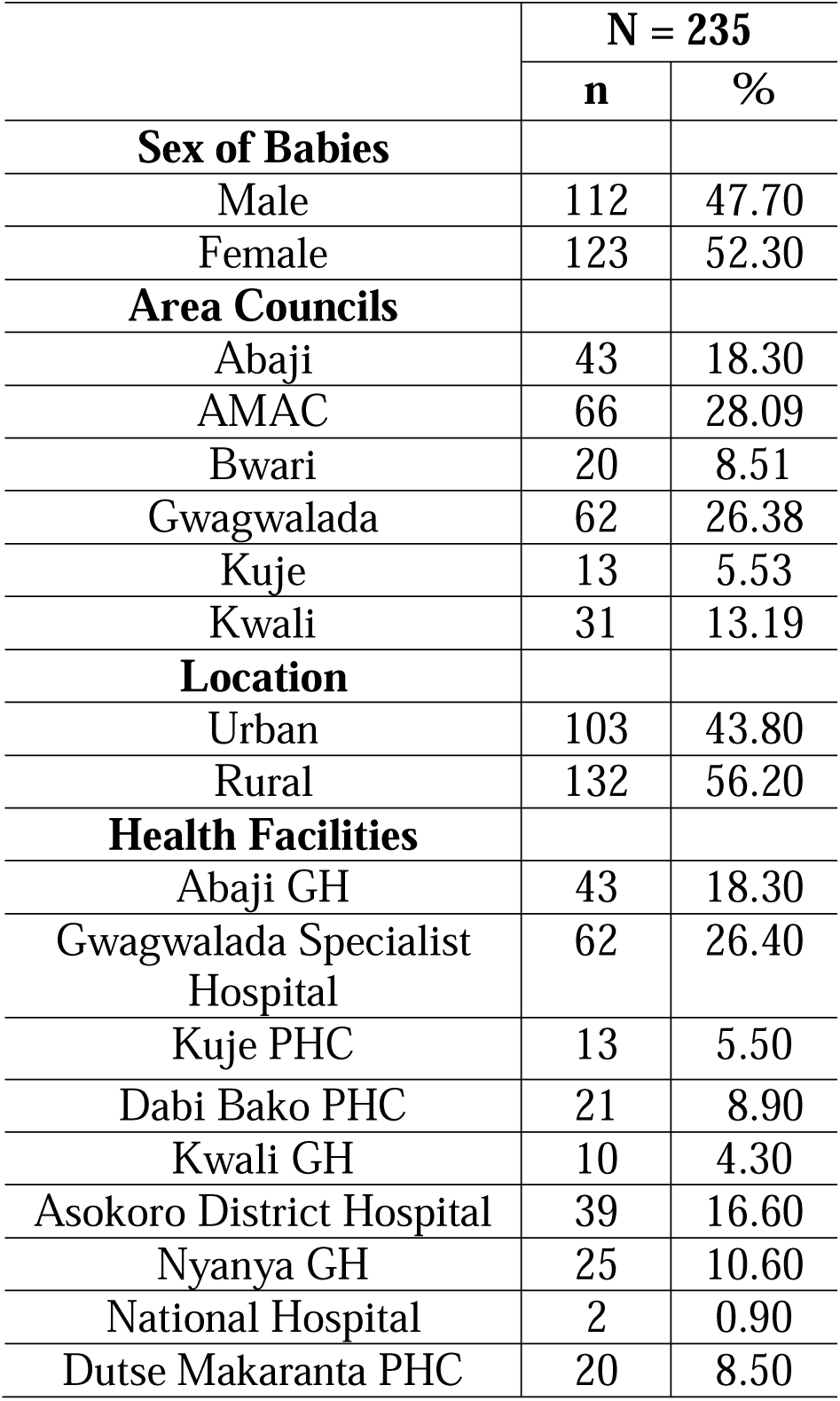
Demographic Information of Recruited Babies for Pentavalent Vaccine Monitoring.

|  | **N = 235** |  |
| --- | --- | --- |
|  | **n** | **%** |
| **Sex of Babies** |  |  |
| Male | 112 | 47.70 |
| Female | 123 | 52.30 |
| **Area Councils** |  |  |
| Abaji | 43 | 18.30 |
| AMAC | 66 | 28.09 |
| Bwari | 20 | 8.51 |
| Gwagwalada | 62 | 26.38 |
| Kuje | 13 | 5.53 |
| Kwali | 31 | 13.19 |
| **Location** |  |  |
| Urban | 103 | 43.80 |
| Rural | 132 | 56.20 |
| **Health Facilities** |  |  |
| Abaji GH | 43 | 18.30 |
| Gwagwalada Specialist Hospital | 62 | 26.40 |
| Kuje PHC | 13 | 5.50 |
| Dabi Bako PHC | 21 | 8.90 |
| Kwali GH | 10 | 4.30 |
| Asokoro District Hospital | 39 | 16.60 |
| Nyanya GH | 25 | 10.60 |
| National Hospital | 2 | 0.90 |
| Dutse Makaranta PHC | 20 | 8.50 |

**Table 2:**
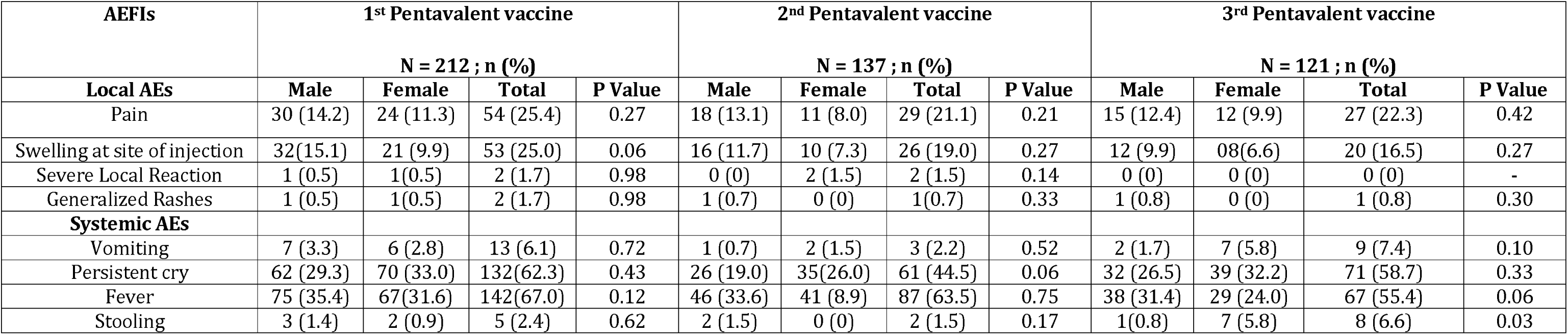
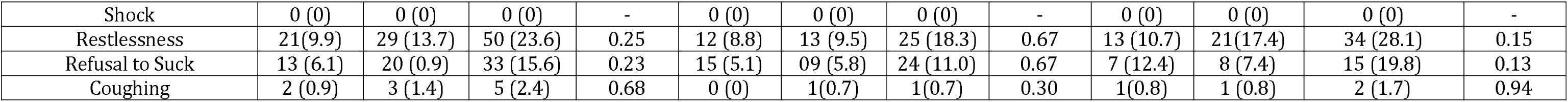
Rates of Local and Systemic Adverse Events Reported from First to Third Pentavalent Vaccination in Recruited Babies.

| AEFIs | 1<sup>st</sup> Pentavalent vaccine |  |  |  | 2<sup>nd</sup> Pentavalent vaccine |  |  |  | 3<sup>rd</sup> Pentavalent vaccine |  |  |  |
| --- | --- | --- | --- | --- | --- | --- | --- | --- | --- | --- | --- | --- |
|  | N = 212 ; n (%) |  |  |  | N = 137 ; n (%) |  |  |  | N = 121 ; n (%) |  |  |  |
| Local AEs | Male | Female | Total | P Value | Male | Female | Total | P Value | Male | Female | Total | P Value |
| Pain | 30 (14.2) | 24 (11.3) | 54 (25.4) | 0.27 | 18 (13.1) | 11 (8.0) | 29 (21.1) | 0.21 | 15 (12.4) | 12 (9.9) | 27 (22.3) | 0.42 |
| Swelling at site of injection | 32 (15.1) | 21 (9.9) | 53 (25.0) | 0.06 | 16 (11.7) | 10 (7.3) | 26 (19.0) | 0.27 | 12 (9.9) | 08 (6.6) | 20 (16.5) | 0.27 |
| Severe Local Reaction | 1 (0.5) | 1 (0.5) | 2 (1.7) | 0.98 | 0 (0) | 2 (1.5) | 2 (1.5) | 0.14 | 0 (0) | 0 (0) | 0 (0) | - |
| Generalized Rashes | 1 (0.5) | 1 (0.5) | 2 (1.7) | 0.98 | 1 (0.7) | 0 (0) | 1 (0.7) | 0.33 | 1 (0.8) | 0 (0) | 1 (0.8) | 0.30 |
| **Systemic AEs** |  |  |  |  |  |  |  |  |  |  |  |  |
| Vomiting | 7 (3.3) | 6 (2.8) | 13 (6.1) | 0.72 | 1 (0.7) | 2 (1.5) | 3 (2.2) | 0.52 | 2 (1.7) | 7 (5.8) | 9 (7.4) | 0.10 |
| Persistent cry | 62 (29.3) | 70 (33.0) | 132 (62.3) | 0.43 | 26 (19.0) | 35 (26.0) | 61 (44.5) | 0.06 | 32 (26.5) | 39 (32.2) | 71 (58.7) | 0.33 |
| Fever | 75 (35.4) | 67 (31.6) | 142 (67.0) | 0.12 | 46 (33.6) | 41 (8.9) | 87 (63.5) | 0.75 | 38 (31.4) | 29 (24.0) | 67 (55.4) | 0.06 |
| Stooling | 3 (1.4) | 2 (0.9) | 5 (2.4) | 0.62 | 2 (1.5) | 0 (0) | 2 (1.5) | 0.17 | 1 (0.8) | 7 (5.8) | 8 (6.6) | 0.03 |
| Shock | 0 (0) | 0 (0) | 0 (0) | - | 0 (0) | 0 (0) | 0 (0) | - | 0 (0) | 0 (0) | 0 (0) | - |
| Restlessness | 21(9.9) | 29 (13.7) | 50 (23.6) | 0.25 | 12 (8.8) | 13 (9.5) | 25 (18.3) | 0.67 | 13 (10.7) | 21(17.4) | 34 (28.1) | 0.15 |
| Refusal to Suck | 13 (6.1) | 20 (0.9) | 33 (15.6) | 0.23 | 15 (5.1) | 09 (5.8) | 24 (11.0) | 0.67 | 7 (12.4) | 8 (7.4) | 15 (19.8) | 0.13 |
| Coughing | 2 (0.9) | 3 (1.4) | 5 (2.4) | 0.68 | 0 (0) | 1(0.7) | 1(0.7) | 0.30 | 1(0.8) | 1 (0.8) | 2 (1.7) | 0.94 |

## 4. Discussion

The widespread adoption of combined or multivalent vaccines in various countries’ immunization schedules has been attributed to the enhanced compliance they offer. While numerous studies have investigated the safety of individual (monovalent) vaccines, it is equally crucial to demonstrate that combined or multivalent vaccines do not adversely affect each other’s reactogenicity [17]. With the continual introduction of new vaccines into national immunization schedules worldwide, the imperative for ongoing monitoring and reporting of both serious and non-serious Adverse Events Following Immunization (AEFIs) cannot be overstated. This vigilance serves to effectively track vaccine safety, enabling informed regulatory decisions by health authorities [8, 9, 11, 12]. By maintaining a robust surveillance system for vaccine safety, countries can uphold public trust in immunization programs and ensure the continued success of efforts to protect population health. This proactive approach not only safeguards individual well-being but also contributes to the broader goals of disease prevention and control on a global scale [6, 8, 9, 11, 12, 13]. The reinforcement of pharmacovigilance systems has emerged as a critical necessity, particularly in low and medium-income countries, with the introduction of new vaccines and vaccine candidates. Many clinical trials have not encompassed these populations, underscoring the importance of robust surveillance mechanisms to ensure vaccine safety [18–25]. Numerous studies have examined the safety of pentavalent vaccines, consistently affirming their tolerable safety profile. Experiences from the initial rollout of pentavalent vaccines in Sri Lanka, Bhutan, India, and Vietnam underscore the pivotal role of enhanced pharmacovigilance systems in monitoring vaccine safety. Through diligent monitoring, these systems assured the safety of the populace and facilitated informed regulatory decisions, ultimately leading to the reinstatement of the suspended pentavalent vaccine.

Despite extensive research on the individual antigens used in pentavalent vaccines, both pre- and post-licensure, it remains imperative to investigate the potential for new and more serious adverse events arising from the combination of components [27]. Sreelakshmi *et al.* [28] highlighted this concern in their study, emphasizing that while identified Adverse Events Following Immunization (AEFIs) may not necessarily be attributable to the pentavalent vaccine, the possibility of unforeseen secondary effects cannot be discounted. They advocated for continuous monitoring of the vaccine’s safety across diverse environments to comprehensively ascertain its true safety and tolerability [21–25]. This ongoing vigilance and research efforts are crucial for maintaining public confidence in vaccination programs and ensuring the continued success of global immunization initiatives. By proactively addressing safety concerns and enhancing pharmacovigilance practices, countries can safeguard public health and effectively mitigate potential risks associated with vaccine administration.[29].

In accordance with this imperative, the present study meticulously assessed the safety and tolerability profile of the combined pentavalent vaccine integrated into Nigeria’s routine immunization schedule. By closely monitoring adverse events following immunization (AEFIs) among infants receiving the vaccine at 6, 10, and 14 weeks, we aimed to provide comprehensive insights into its safety profile [20, 21, 22]. The most frequently reported local AEFIs during the study period were pain and swelling at the injection site, while fever emerged as the predominant systemic adverse event, occurring in 66.98% of cases. These findings align closely with studies by Angela *et al.* [31] and Sharma *et al.* [33]. Furthermore, our investigation revealed no serious AEFIs necessitating hospital visits. The identified adverse events were predominantly non-serious and typically resolved either with analgesics or without medication within 48 hours post-immunization. These results are consistent with findings from several other studies on pentavalent vaccines conducted by Bavdekar *et al.* [30], Dodoo *et al.* [32], Asturias *et al.* [34], and Manoochehr *et al.* [36] (2017).

Moreover, our study demonstrated a decline in the frequency of adverse events with subsequent vaccinations, indicating an acceptable safety profile over the vaccination series. This observation is corroborated by the study conducted by Eregowda *et al.* [29]. Overall, our findings affirm that the pentavalent vaccine is well tolerated and safe when administered to infants in the Federal Capital Territory (FCT), Nigeria, as per the approved immunization schedule. Notably, the sex of the infants did not exhibit statistically significant differences in the types or occurrence of reported AEFIs during our study. The consolidation of five different antigens into a single vaccine shot has significantly bolstered protection against five distinct vaccine-preventable diseases among infants, facilitating enhanced compliance and vaccine uptake. Consequently, this integrated approach has contributed to a marked reduction in the prevalence of these diseases among infants in Nigeria, underscoring the pivotal role of the pentavalent vaccine in public health initiatives.

## 5 Conclusion

The introduction of the pentavalent vaccine into Nigeria’s routine immunization schedule has proven to be both safe and well-tolerated for infants aged 6 to 14 weeks. This comprehensive study aimed to assess the vaccine’s safety profile by closely monitoring adverse events following immunization (AEFIs). The findings indicate that the vaccine is effective and poses minimal risk to the infants, providing reassurance to healthcare providers and parents alike. The study revealed that the AEFIs reported were predominantly non-serious. These included local reactions such as pain and swelling at the injection site, and systemic reactions such as fever. Crucially, these adverse events resolved within 48 hours either with or without medication. The absence of serious AEFIs requiring hospitalization underscores the vaccine’s safety and supports its continued use in routine immunization programs.

Furthermore, the analysis showed no statistically significant differences in the occurrence of AEFIs between male and female infants. This finding highlights the vaccine’s consistent safety and tolerability across different genders, reinforcing its suitability for widespread use. The ability to protect against five major childhood diseases with a single vaccine shot simplifies the immunization process and promotes higher compliance among parents. Overall, the introduction of the pentavalent vaccine in Nigeria represents a significant advancement in public health. By providing comprehensive protection against multiple diseases, the vaccine plays a crucial role in reducing the prevalence of these conditions among infants. The study’s positive findings support the continued inclusion of the pentavalent vaccine in Nigeria’s immunization schedule, ensuring that infants receive safe and effective protection from an early age.

## 6. Study Limitations

Despite the promising findings of this study, several limitations must be acknowledged. First, the study relied on self-reported data from mothers, which can be subject to recall bias and inaccuracies. Although efforts were made to train the mothers on proper recording techniques, the potential for human error in documenting adverse events cannot be completely eliminated. Second, the study’s sample size, while sufficient for initial safety assessments, may not be large enough to detect very rare adverse events. Larger, multi-center studies would be necessary to identify less common reactions and provide a more comprehensive safety profile. Additionally, the study was conducted in a limited geographical area, the Federal Capital Territory (FCT) of Nigeria, which may not be fully representative of the diverse populations and healthcare settings across the country. Another limitation is the relatively short follow-up period of three months. While this timeframe is adequate for identifying immediate and short-term AEFIs, it does not capture long-term safety outcomes. Longer follow-up periods are needed to monitor for any delayed adverse events and to better understand the full impact of the pentavalent vaccine. Lastly, the study did not include a control group of unvaccinated infants or infants receiving different vaccines for comparative analysis. This makes it difficult to attribute the observed AEFIs solely to the pentavalent vaccine without considering other potential confounding factors. Future studies should incorporate control groups to strengthen the causal inferences regarding vaccine safety.

## 7. Significance Statement

The introduction of the pentavalent vaccine into Nigeria’s routine immunization program marks a significant milestone in public health, offering protection against five major childhood diseases in a single shot. This study reinforces the vaccine’s safety and tolerability, demonstrating that adverse events following immunization are predominantly non-serious and self-limiting. These findings provide critical support for the ongoing use of the pentavalent vaccine in Nigeria and potentially other similar settings. By confirming the safety of the pentavalent vaccine, this study helps to build trust among parents and caregivers, which is crucial for maintaining high vaccination rates. High compliance with immunization schedules is essential for achieving herd immunity and reducing the prevalence of vaccine-preventable diseases. The study’s results underscore the importance of robust pharmacovigilance systems in ensuring vaccine safety and efficacy. Moreover, the absence of significant differences in adverse event occurrences between male and female infants highlights the vaccine’s consistent performance across different demographic groups. This uniform safety profile supports the equitable use of the pentavalent vaccine, ensuring that all infants, regardless of gender, receive the same level of protection against debilitating diseases. In summary, this study not only validates the safety of the pentavalent vaccine but also underscores its critical role in enhancing public health outcomes. The findings advocate for the continued integration of the pentavalent vaccine into routine immunization programs, ultimately contributing to the global efforts in reducing childhood morbidity and mortality from preventable diseases. The study’s insights are invaluable for policymakers, healthcare providers, and public health advocates working to improve immunization coverage and safeguard children’s health.

## Data Availability

All data produced in the present work are contained in the manuscript

## Acknowledgments

The authors would like to express their heartfelt gratitude to all the participants who contributed to this study. Your willingness to share your time and responses has been invaluable to the success of this research. We deeply appreciate your participation and the insights you provided, which have significantly enriched the findings of this study. Thank you for your essential role and commitment, which made this work possible.

## Author contributions

All authors collaboratively developed the concept and design of the study. They were collectively involved in data collection and analysis, ensuring a comprehensive approach to the research. Each author contributed to drafting the initial article, bringing their unique perspectives and expertise to the writing process. Furthermore, all authors participated in interpreting the data, providing critical insights that shaped the study’s conclusions. They thoroughly reviewed the content of the article, ensuring accuracy and coherence, and ultimately approved the final version for publication. This collaborative effort highlights the integral contributions of each author at every stage of the research.

## Declarations

Approval for the study was obtained from the Ethics Committee of the Federal Capital Territory Health Ethics Committee, with Protocol Approval Number: **FHREC/2015/01/62/21-10-15**. Administrative approvals were also secured from the Federal Capital Territory Development, FCT Primary Health Care Board, and Asokoro District Hospital, FCT. Prior to participation, all study participants were provided with oral explanations regarding the study’s objectives and the assurance of anonymity. Informed consent was obtained from each participant before the commencement of the interview process. Confidentiality of all collected data was strictly maintained throughout the study duration, ensuring the privacy and integrity of participants’ information.

## Consent for publication

Not applicable.

